# Ultra-fast one-step RT-PCR protocol for the detection of SARS-CoV-2

**DOI:** 10.1101/2020.06.25.20137398

**Authors:** Ehsan Asghari, Anna Höving, Paula van Heijningen, Annika Kiel, Angela Krahlemann-Köhler, Melanie Lütkemeyer, Jonathan Storm, Tanja Vollmer, Cornelius Knabbe, Barbara Kaltschmidt, Gert de Vos, Christian Kaltschmidt

## Abstract

The COVID-19 pandemic resulted in lockdowns all over the world thus affecting nearly all aspects of social life and also had a huge impact on global economies. Since vaccines and therapies are still not available for the population, prevention becomes desperately needed. One important aspect for prevention is the identification and subsequent isolation of contagious specimens. The currently used methods for diagnostics are time consuming and also hindered by the limited availability of reagents and reaction costs, thus presenting a bottle neck for prevention of COVID-19 spread. Here, we present a new ultra-fast test method which is ten times faster than conventional diagnostic tests using real time quantitative PCR (RT-qPCR). In addition, this ultra-fast method is easy to handle as well as cost effective. We translated published SARS-CoV-2 testing protocols from the Centers of Disease Control and Prevention (Atlanta, Georgia, USA) and the Charité Berlin (Germany) to the NEXTGENPCR (NGPCR) machine and combined it with a fluorescence-based endpoint measurement. Fluorescence was measured with a commercial blue light scanner. We confirmed the NEXTGENPCR results with commercially available positive controls. In addition, we isolated RNA from SARS-CoV-2 infected patients and achieved similar results to clinical RT-qPCR assays. Here, we could show correlation between the results obtained by NEXTGENPCR and conventional RT-qPCR.

## Introduction

The COVID-19 pandemic of late 2019 is a major threat to people’s health worldwide, with no approved therapeutic drugs or vaccines available^1^. Counting 7,924,572 confirmed cases and 434,367 deaths worldwide by the 16^th^ of June 2020^2^, most common measures to slow down the spread of the virus are approaches of limiting social contacts including lockdowns and quarantines. These measures, designed to keep healthcare systems from collapsing, have huge impacts on social life and economies^3^. Infectious specimens are not always perceivable, as symptoms might be light in many cases^4^, thus loosening social life restrictions might lead to a second wave of infection. The great initial success of large-scale testing in South Korea shows that testing capacity can be a key in controlling outbreaks^5^. The most prevalent testing method for the SARS-CoV-2 virus currently is the use of one-step reverse transcription quantitative polymerase chain reaction (RT-qPCR)^6,7^. Despite RT-qPCR being a generally well-established diagnostic technique^8^, testing for SARS-CoV-2 is currently limited in many regions^9^. Two current issues limiting the testing capacities are the availability of the reagents^10^ and the technical capability of the qPCR machines limiting the total amount of possible tests per day.

The NEXTGENPCR (NGPCR) machine is a new platform capable of performing polymerase chain reactions in short time frames^11^. A mechanical system moves a reaction plate between three zones of heating at different temperatures. Each zone consists of two blocks at equal temperature. Samples are contained in thin plastic foils, which are pressed between the two perpendicular blocks in a zone, ensuring sample mixing. This allows instant temperature changes of the sample, thus removing the limitations of classical Peltier element-based PCR machines. The reaction is no longer slowed down by the ramping rates at different temperatures. The NEXTGENPCR machine combines ultrafast cycling with the possibility to perform a RT-step within the same program, before the PCR reaction.

In this study, we adapted two commonly used SARS-CoV-2 diagnosis protocols for usage with the NEXTGENPCR machine. Both assays use hydrolysis probes (ex. TaqMan), making them suitable for fluorescence-based endpoint measurements to determine amplification. The primer/probe sets for the detection of the viral nucleocapsid 1 and 2 (N1 and N2) genes from “Centers for Disease Control and Prevention (CDC) 2019-Novel Coronavirus (2019-nCoV) Real-Time Reverse Transcriptase (RT)-PCR Diagnostic Panel” (in the following: CDC panel)^12^ were used together with the corresponding positive control DNA plasmid^6^. Furthermore the primers and the probe for the E gene of the SARS-CoV-2 virus from the protocol of the Charité Berlin (Prof. Drosten, “The Berlin Panel”, in the following: Drosten panel) were used^7^. These widely used protocols however present only an example, as most RT-qPCR based methods should be adaptable as well.

## Material and Methods

### Collection and preparation of samples

RNA from respiratory samples was obtained from the Institut für Laboratoriumsund Transfusionsmedizin, Herzund Diabeteszentrum NRW (HDZ), Universitätsklinik der Ruhr-Universität Bochum, Bad Oeynhausen, Germany. Information of the COVID-19 tested patients is summarized in Table 1. All samples were collected by the authors in accordance with the German Act on Medical Devices (MPG, guideline 98/79/EG) for the collection of human residual material to evaluate suitability of an in vitro diagnostic medical device (§24). The need for informed consent and ethical approval was waived since all materials used were anonymous samples already analysed in routine laboratory diagnostics. Extraction of total RNA from throat swab samples was done fully automated using the AltoStar Automation System (AM16, Altona Diagnostic Technologies (ADT), Hamburg, Germany) according to the manufacturer’s instructions. Swabs were squeezed in 800 µl phosphate buffered saline (PBS), 500 µl sample was used for RNA extraction. RNA was eluted in 45 µl elution buffer. Amplification using the RealStar SARS-COV-2 RT-PCR assay 1.0 (ADT) was performed according to the manufacturer’s instructions using 10 µl eluate combined with 20 µl of mastermix on the Bio-Rad CFX 96 Deep Well Real-time PCR detection system (part of the AM16 system, ADT).

**Table 1:** Basic information for the clinical samples collected from the COVID-19 tested patients.

| Patient | Sex | Age range | CT value AM16 |
| --- | --- | --- | --- |
| 1 | w | teens | 23.57 |
| 2 | w | teens | 27.33 |
| 3 | w | 50s | 22.29 |
| 4 | m | teens | 31.27 |
| 5 | m | 60s | 19.06 |
| 6 | w | 20s | 29.54 |
| 7 | m | 50s | 17.83 |
| 8 | m | 50s | 26.53 |
| 9 | m | 20s | 20.84 |
| 10 | w | 70s | 27.96 |

### Preparation of positive controls

2019-nCoV_N_Positive Control from IDT Leuven, Belgium was used as a positive control for the CDC 2019-Novel Coronavirus (2019-nCoV) Real-Time RT-PCR diagnostic panel in the recommended concentration of 1000 copies per reaction. Twist Synthetic SARS-CoV-2 RNA control 1 from Twist Biosciences San Francisco, USA was used in a 10 fold dilution series from 100.000 copies per reaction as positive RNA control.

### Selection of the primer pairs and probes

For the COVID-19 test following the CDC 2019-Novel Coronavirus (2019-nCoV) Real-Time RT-PCR diagnostic panel the PCR test Kit from IDT Leuven, Belgium: 2019-nCoV CDC EUA was used. The kits include primers and 5’ FAM / 3’ Black Hole Quencher® (BHQ) probes (listed in Table 2). Additionally, for the COVID-19 test following the protocol of Charité Berlin (Prof. Drosten, “The Berlin Panel”) the PCR test Kit from IDT Leven, Belgium was used. The Drosten panel detects the E gene to positively identify SARS-CoV-2. The kits include primers for the E gene and 5’ FAM-ZEN™/Iowa Black® FQ probes (listed in Table 3).

**Table 2:** Primer/probe pairs for the CDC 2019-Novel Coronavirus (2019-nCoV) Real-Time RT-PCR diagnostic panel.

| Reagent Label | Description | Quantity/Tube | Reactions/Tube |
| --- | --- | --- | --- |
| 2019-nCoV_N1 | 2019-nCoV_N1<br>Combined Primer/Probe<br>Mix | 22.5 nmol | 1000 |
| 2019-nCoV_N2 | 2019-nCoV_N2<br>Combined Primer/Probe<br>Mix | 22.5 nmol | 1000 |

**Table 3:**
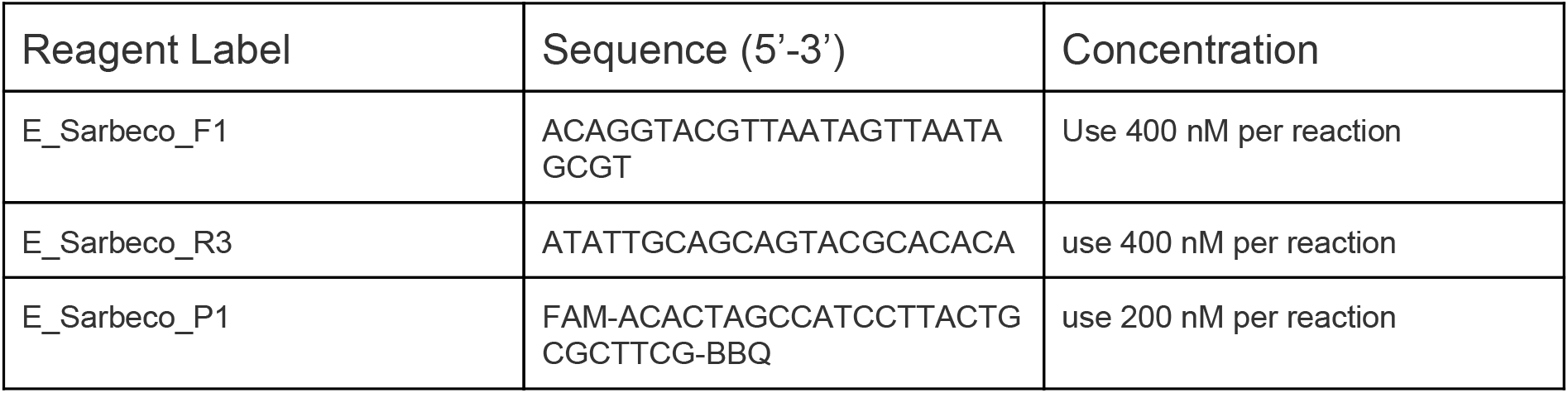
Primer/probe pairs for the Drosten protocol for 2019-Novel Coronavirus (2019-nCoV) Real-Time RT-PCR diagnostic panel.

### PCR Assays

Each clinical RNA sample assayed in-house was run in duplicate using 1 µl of the eluate as a template. Direct reverse transcription with the qScript™ XLT ToughMix® from Quanta Biosciences (Beverly, Mass., USA) was performed. As incubation time for the RT-step, 5 min at 55 °C was used in the NEXTGENPCR machines and 10 min in the RT-qPCR machine. The reagent volumes in the CDC Assay and the Drosten Assay were used as instructed by the manufacturers. The optimized protocol used for the NEXTGENPCR is shown in Fig. 2 F. For the RT-qPCR we used a Corbett Rotor-Gene 6000 (Qiagen, Hilden, Germany) with the settings shown in Table 4.

**Table 4:** Protocol for RT-qPCR using the Corbett Rotor-gene 6000

| Temp. (°C) | Duration (s) | Repeat |
| --- | --- | --- |
| 55 | 600 | 1 |
| 95 | 120 | 1 |
| 95 | 15 | 45 |
| 55 | 30 | 45 |
| 72 | 20 | 45 |

## NEXTGENPCR

Plates from the NEXTGENPCR machine contained the same assay, producing a fluorescent signal with positive samples. The fluorescence of the wells from the NEXTGENPCR plates were measured with a fluorescence scanner (MicroTek Bio1000F, MBS). NEXTGENPCR QuickDetect™ software was used to detect the liquid within each well of the plate and score the fluorescence levels.

For each assay, additional plates with no template controls (NTCs) were prepared and the mean of the NTCs plus three times the standard deviation was set as the threshold for quantification. Absolute fluorescence values measure from 0 to 256. If only one of the measured wells showed a grey level above the threshold level, the value of the well qualifying as negative was excluded from further analysis and the sample was called positive. If both wells showed grey levels below the threshold value, the sample was called negative and no quantification value was shown in the quantification graphs.

## Results and Discussion

To visualize the workflow used in this study we made a graphical abstract (Fig 1). In Fig. 1 A the NEXTGENPCR machine is shown. NEXTGENPCR uses ultra-thin SBS formatted plates which are available in 96 well for 5 and 20 µl and in 384 well for 5 µl (details at www.nextgenpcr.com). The plates are heat sealed with transparent seals. For measuring the hydrolysed fluorescent probes, a fluorescence scanner (MicroTek Bio1000F) obtained from MBS was used (Fig. 1 C). PCR plates were scanned (Fig. 1 D) and quantified with QuickDetect™ (Fig. 1 E), which automatically recognizes the liquid within each well and quantifies fluorescence in 256 grey levels. The software scores each well, positive (red) or negative (green), based on user-defined threshold values. A typical result of the quantification is shown in Fig. 1 F.

**Figure 1:**
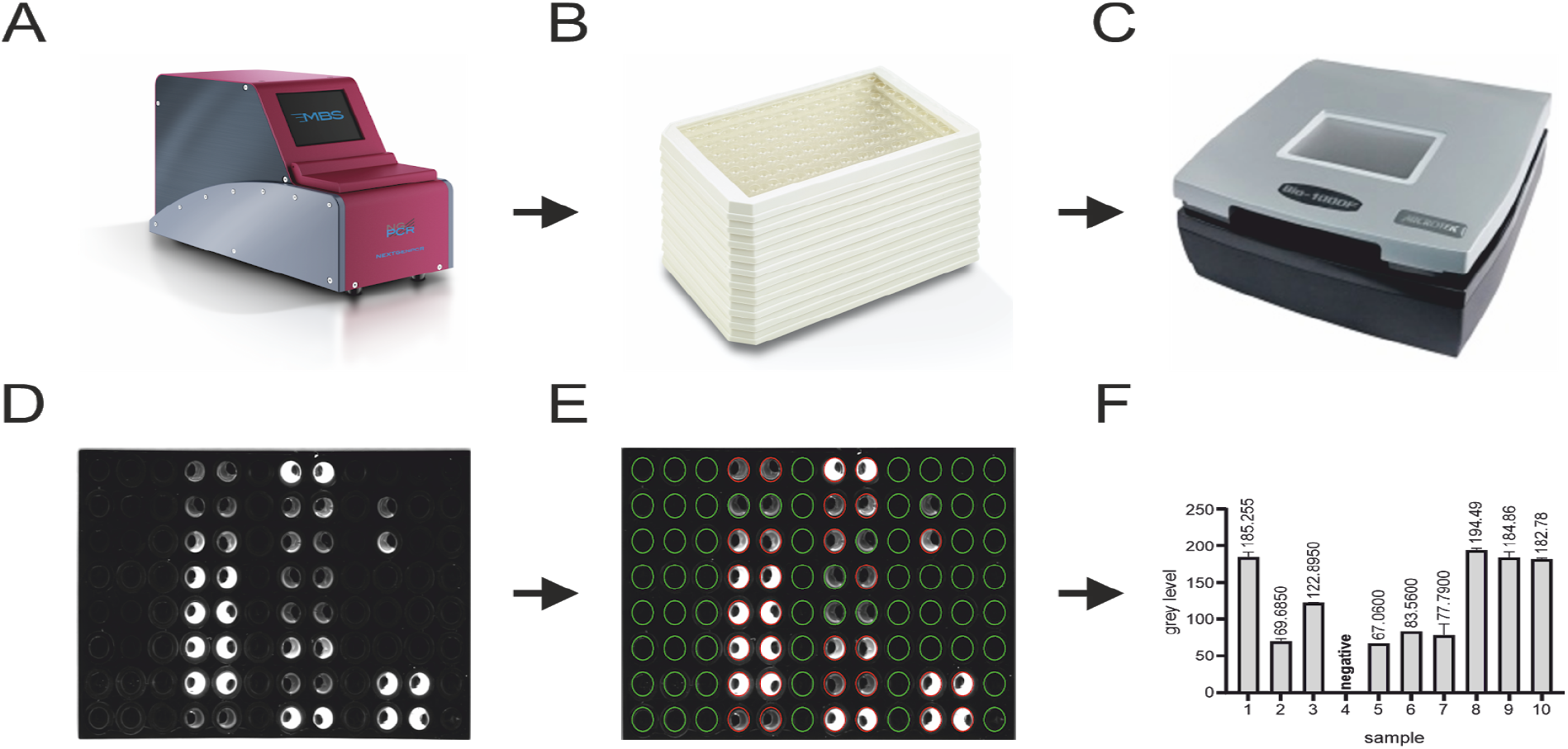
Graphical workflow: A PCR reaction is performed on the NEXTGENPCR Machine **(A)**. The plate containing the wells with the reaction **(B)** is imaged using a bluelight reader **(C)**. The image **(D)** is measured using the quantification software. The wells with fluorescence values below a set threshold level are marked in green, the wells above the threshold are marked in red **(E)**. The measured fluorescence values can also be shown as bar chart **(F)**.

We performed an optimization in the NEXTGENPCR machine of the CDC panel protocol using the according plasmid standard (Fig. 2 A, B, C). We determined that the protocols using 35 and 40 cycles are sufficient to detect at least 2000 plasmid copies, see figure 2 b and c. 40 and 45 cycles were efficient in amplification of as low as 40 copies. However, at the end of 45 cycles, some NTC controls appeared positive. We therefore conclude that limitation of cycle number to 40 in endpoint PCR might be a way to avoid false positive results. In these experiments with 40 cycles PCR amplification time was 10 min.

**Figure 2:**
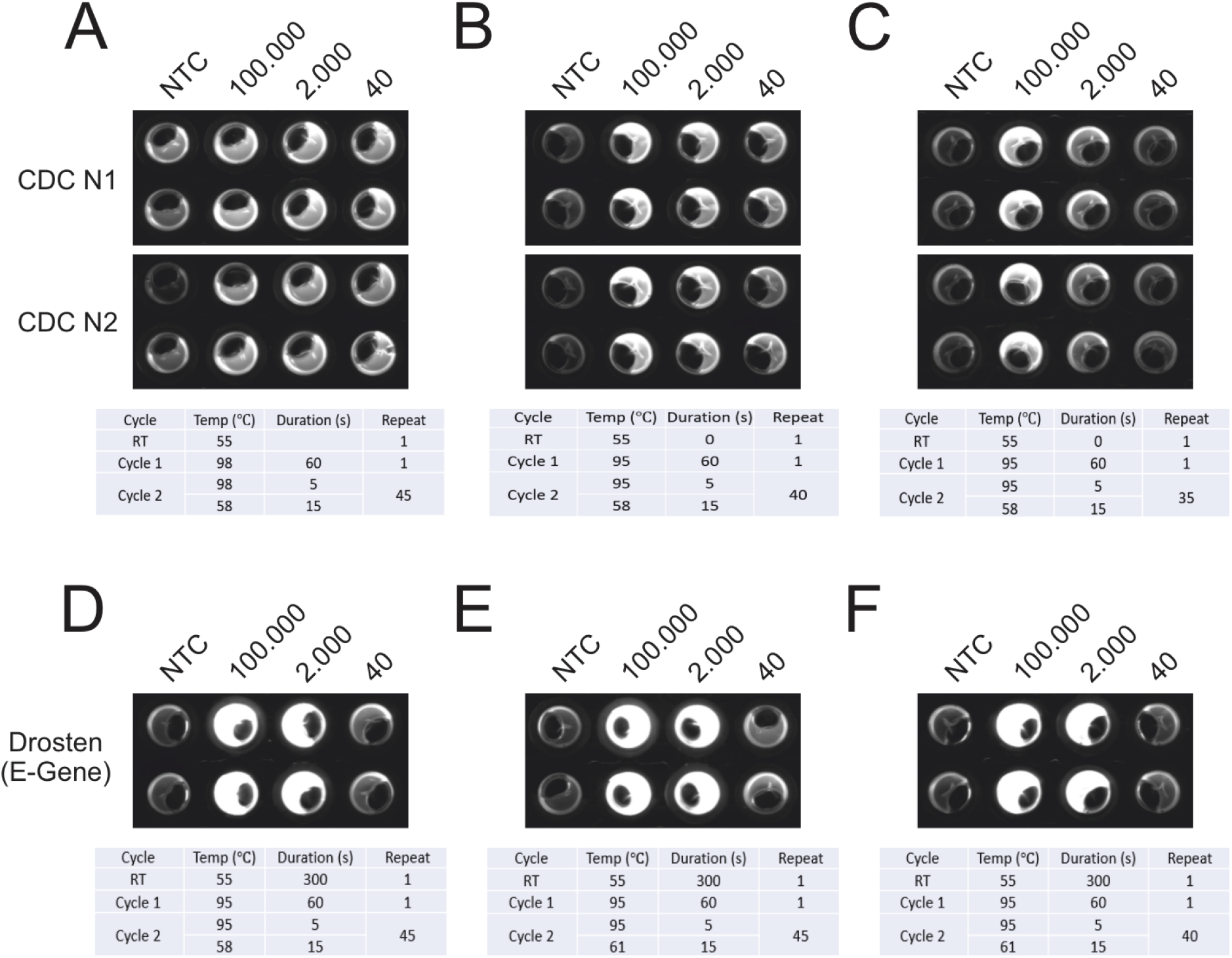
Optimization of different protocols for the NEXTGENPCR: The CDC panel **(A-C)** and the Drosten panel **(D-F)** were ran at different settings to determine optimal conditions. The CDC panel was performed using 45 **(A)**, 40 **(B)** and 35 **(C)** cycles. The Drosten panel was ran at an elongation temperature from 58 °C **(D)** and 61 °C **(E, F)** and with 45 **(D, E)** and 40 **(F)** cycles.

The optimization of the Drosten panel was performed using synthetic RNA (Twist Synthetic SARS-CoV-2 RNA Control 1, Twist Bioscience San Francisco), thus in contrast to the CDC panel optimization included an RT step. Comparison between annealing at 58 °C (Fig.2 D) and 61 °C (Fig.2 E) yielded no obvious difference in amplification efficiency. Like 45 cycles (Fig.2 D, E), 40 cycles (Fig.2 F) lead to comparable results. As a positive side effect, the limitation of cycles can avoid false positive results as shown above and is considerably time saving as well. Thus, the protocol of Fig. 2 F was chosen for further experiments. Taken together, total reaction time including RT and PCR amounts to 16 min using 40 cycles. According to a press release from GNA biosolutions (published at munich-startup.de), their SARS-CoV-2 test in development might take 15 min. We see an advantage in the setup used here, since standard primer panels can be used and no dedicated gold nanoparticle-coupled probes are necessary.

Additionally, we tested the reproducibility of the protocol used in Fig. 2 C on five different NEXTGENPCR machines (Fig. 3 A-E). We were able to reproduce identical results on all five machines. Furthermore, we excluded the occurrence of edge effects by testing different positions on the plate (Fig. 3 F) using the protocol shown in Fig.2 F. Taken together, the reactions seem to be robust and transferable.

**Figure 3:**
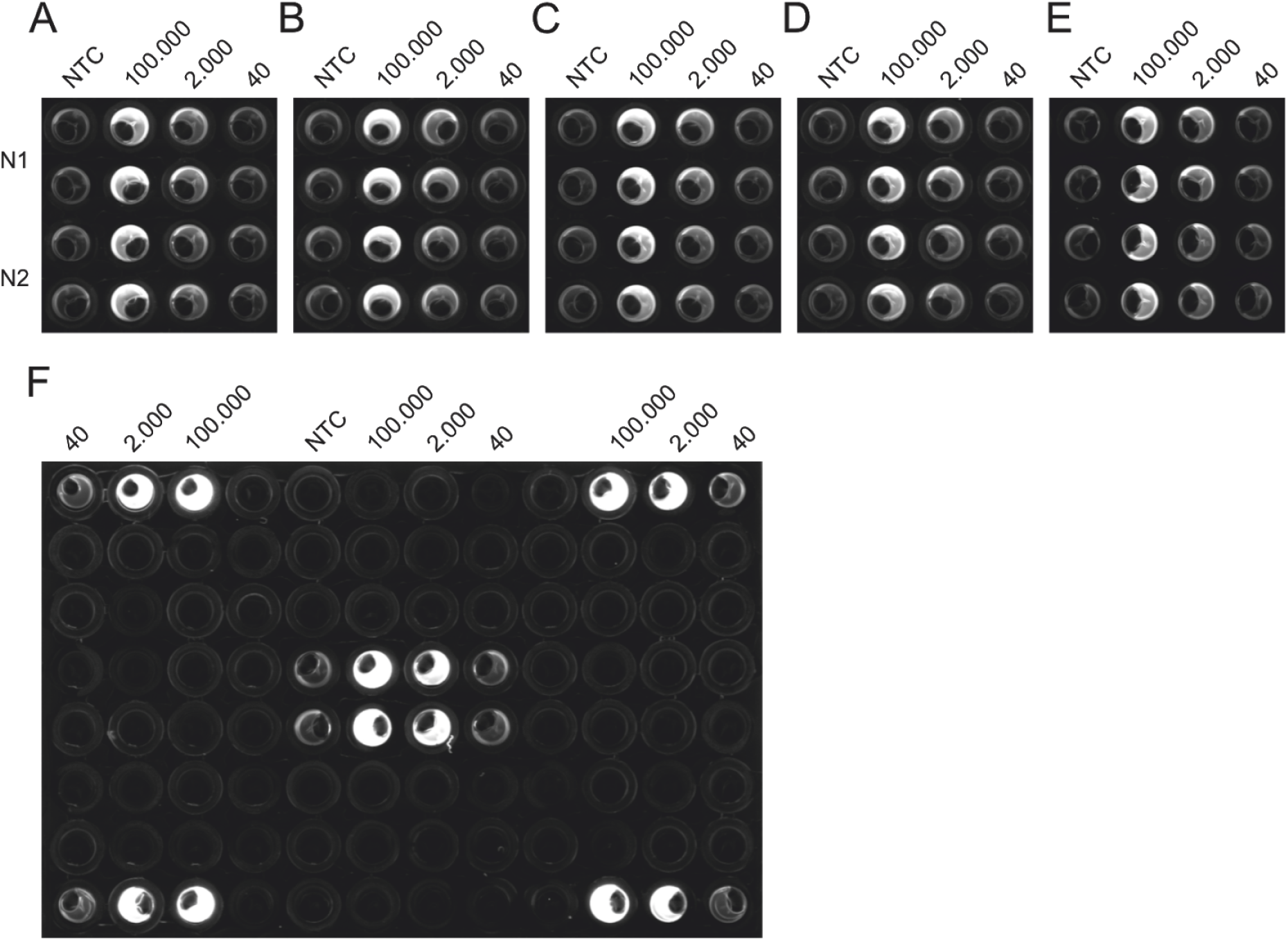
Analysis of reproducibility and equal performance of different PCR machines: The CDC panel was performed on five different machines, using the same master-mixes to test for equal performance **(A-E)**. The Drosten panel was performed on various wells of the same plate to control for edge-effects possibly occurring from impaired heating or measurement based on proximity to the edge of the plate **(F)**.

We obtained RNA eluates from positive samples from an accredited clinical diagnostic laboratory (*HDZ) and performed a RT-qPCR using the Drosten panel to ensure the sample quality was not compromised from storage and transport. Comparison of our results with the Cq values obtained by the RT-qPCR, performed at the clinical diagnostic laboratory using the same primer/probe set, showed a quite similar pattern. Maximal difference amounted to 2.5 cycles in RNA sample 2. We conclude that we could reproduce RT-qPCR results with the protocol established at our lab. Remarkably, we only used one tenth of the sample volume in our reactions as compared to the clinical diagnostic laboratory.

Similar results were obtained with the NEXTGENPCR using the Drosten panel. Importantly, this protocol only takes approximately 10 % of the time necessary for our RT-qPCR protocol. Undoubtedly, the samples 1, 3, 8, 9 and 10 appear positive by the Drosten panel in RT-qPCR as well as NEXTGENPCR. Strikingly these results are directly observable by eye in the endpoint NEXTGENPCR assay (Fig.4 B). Qualitative scoring by visual inspection of the NEXTGENPCR tests becomes difficult with samples that have a low amount of viral RNA (copies with corresponding Cq values above 28; see Fig. 4 samples 2,4,5,6, and 7). QuickDetect™ will also score lower fluorescent values and discriminate between positives and negatives. Thus, we conclude the endpoint-based method is a great simplification, allowing for rapid diagnosis of high load samples.

**Figure 4:**
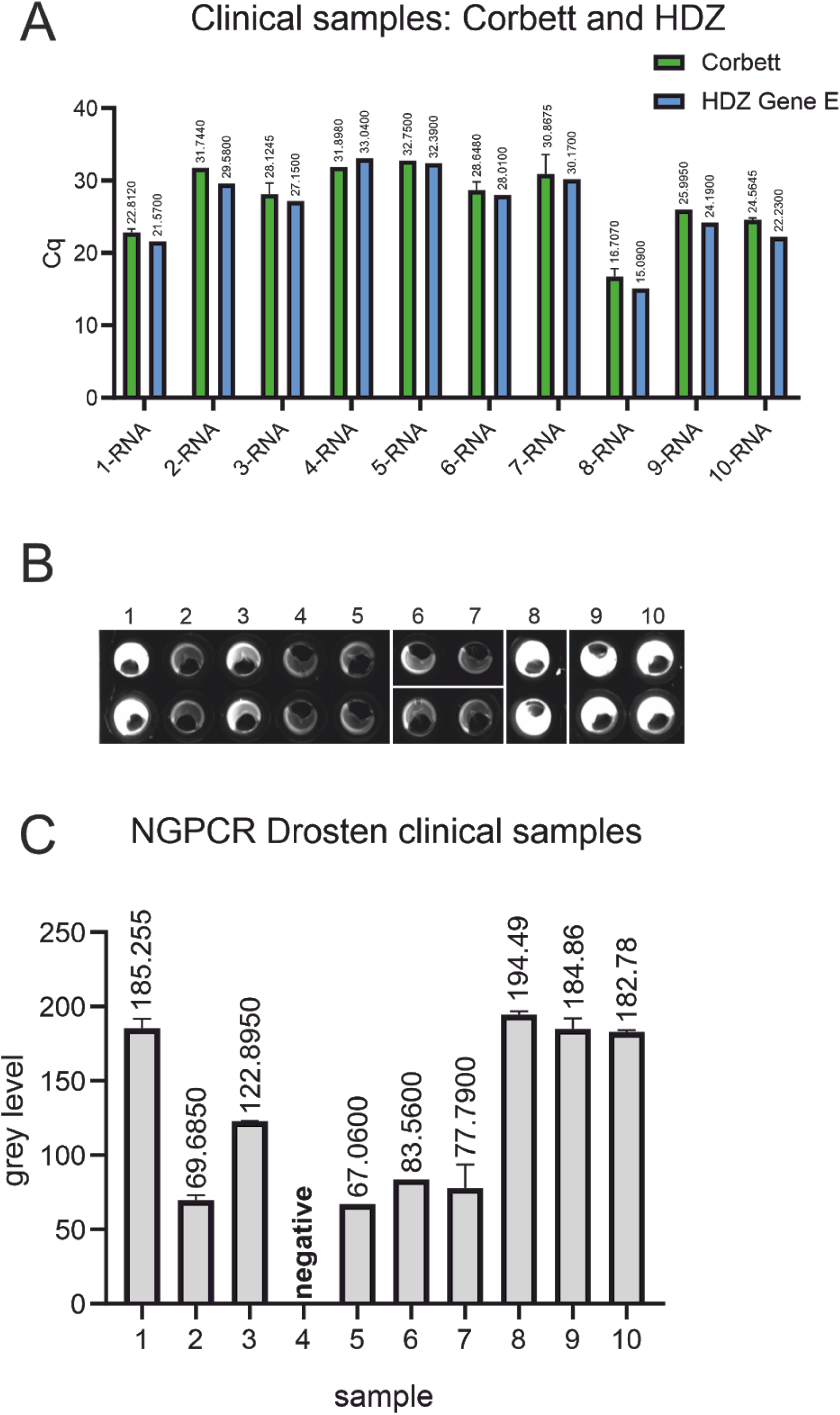
Comparison of Cq values obtained at the HDZ and the laboratory qPCR machine: The RT-qPCR of the obtained RNA from clinical samples was repeated using an altered protocol with a different machine and one tenth of the sample volume used for clinical tests to validate the laboratory assays used **(A)**. The Drosten panel applied was also ran on the NEXTGENPCR and the measured fluorescence **(B)** was quantified and is shown as a bar-plot **(C)**.

The official recommendation for RT-qPCR assay positivity from the CDC are Cq values below 40 cycles^6^, rendering the CDC non template controls positive in our hands (Fig. 5 A). All NTC wells in this plate were called positive using the NEXTGENPCR method (in the range of 1 synthetic RNA-copy). This could be caused by contamination, maybe resulting from the absence of a clean room environment in our assays. The threshold for the NEXTGENPCR could be adjusted accordingly. For the RT-qPCR of the Drosten panel, the threshold for positivity can be set at the recommended 40 cycles. As the CDC N1 assay turned positive in the NTC with a Cq value of 36.5, we suggest a threshold of 36. For the CDC N2 assay we suggest a threshold of 38 cycles based on similar reasoning. The sensitivity of the Drosten- and the CDC RT-qPCR as well as tests performed on the NEXTGENPCR machine ranged between 1 to 10 synthetic RNA copies (Fig. 5).

**Figure 5:**
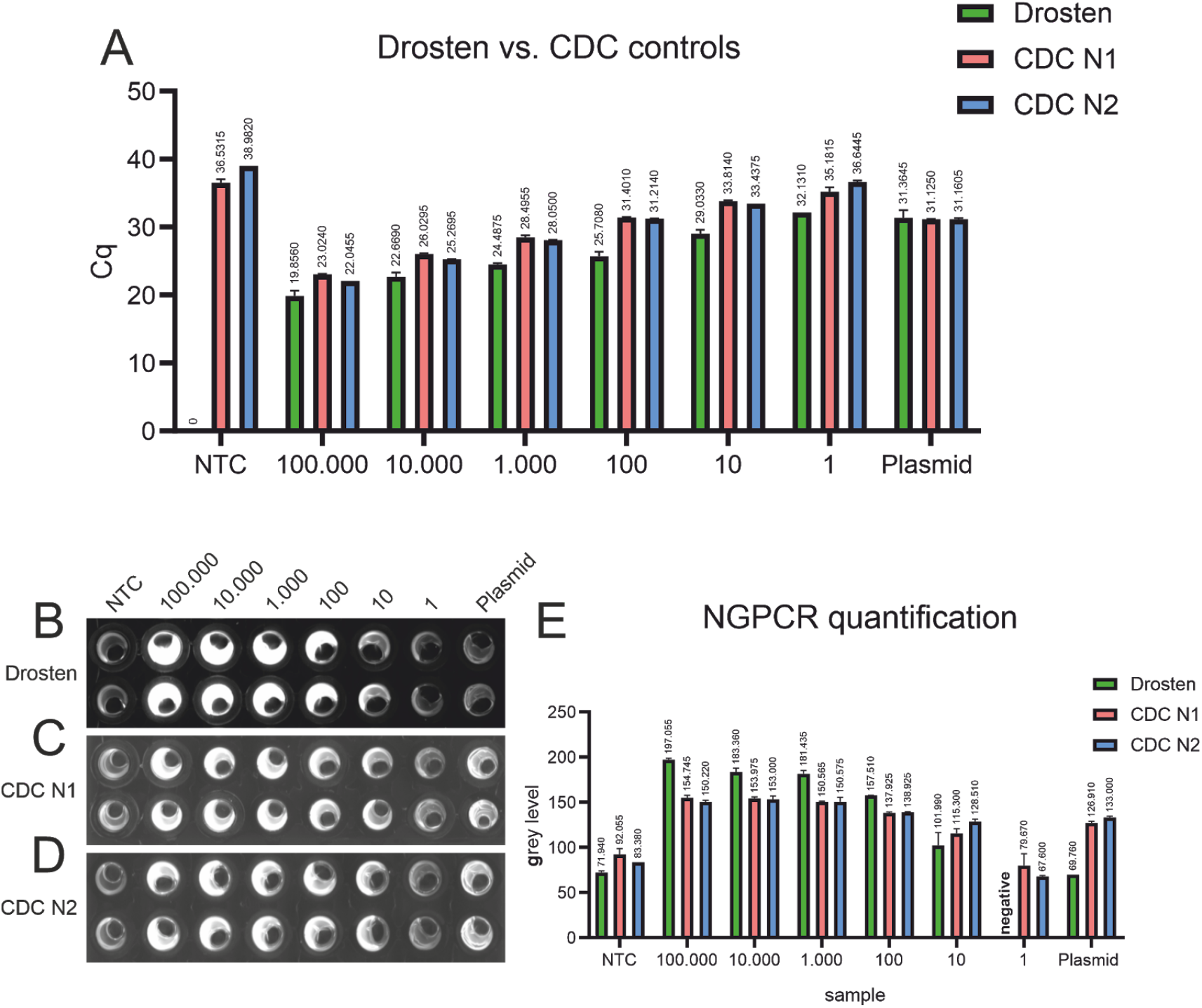
Detection limit of RT-qPCR and NEXTGENPCR: RT-qPCR of synthetic RNA with the Drosten-panel as well as with the CDC N1- and N2-protocols. Cq-values are shown above the respective bars **(A)**. Fluorescence-images after NEXTGENPCR of synthetic RNA with the Drosten-panel as well as with the CDC N1- and N2-protocols **(B, C, D)**. Quantification of NEXTGENPCR of synthetic RNA with the Drosten-panel as well as with the CDC N1- and N2-protocols. Grey levels are shown above the respective bars **(E)**.

Samples were scored positive if either replicate exceeded the thresholds set according to the values determined in Figure 5. The RT-qPCR assay using the Drosten panel called all of the samples positive (Fig. 6 A), resulting in 100% correlation with the results from an accredited clinical diagnostic laboratory (Fig. 4 A). The NEXTGENPCR method using the Drosten panel resulted in 9 of 10 positive samples being correctly called (sample 4 was called negative). (Fig. 6 B, E). Using the CDC N1 primer/probe pair on the RT-qPCR platform we failed to determine samples 4 and 5 as positive. However, using the CDC N2 assay resulted in 100% correct calls. (Figure 6A) The CDC N1 and N2 assays on NEXTGENPCR resulted in 100% correlation with diagnostic laboratory results (Fig. 6 C, D, E). The discordant samples may have failed to be detected since only 20% of the recommended samples volume was available for the testing here. These samples could not be re-tested due the limited amounts donated for this study.

**Figure 6:**
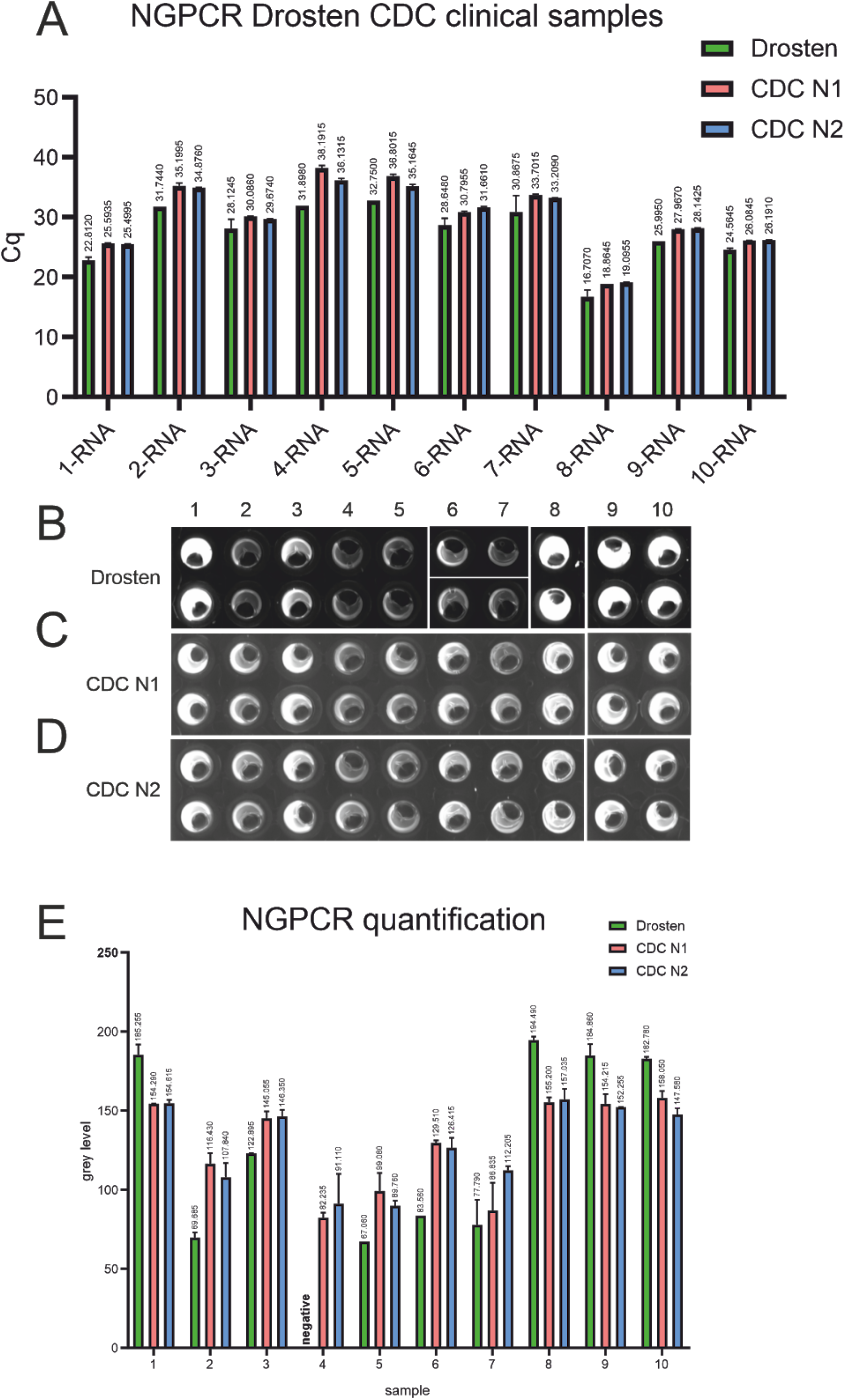
Performance of the CDC- and the Drosten panel in the detection of virus RNA in clinical samples: RT-qPCR of clinical RNA-samples measured with the Drosten-panel as well as with the CDC N1- and N2-protocols. Cq-values are shown above the respective bars **(A)**. Fluorescence image after NEXTGENPCR of of clinical RNA-samples measured with the Drosten-panel as well as with the CDC N1- and N2-protocols **(B, C, D)**. Quantification of NEXTGENPCR results of clinical RNA-samples measured with the Drosten-panel as well as with the CDC N1- and N2-protocols. Grey levels are shown above the respective bars **(E)**.

We aimed to analyse a potential correlation of Cq values measured by RT-qPCR with grey levels measured in a scanner after endpoint PCR using NEXTGENPCR. Therefore, we plotted Cq values on the y-axis and grey levels on the x-axis. Strikingly, we could see a good correlation of the endpoint measurement with the Cq values with a R^2^ of 0.94 for the Drosten panel tested with a synthetic RNA dilution series (Fig. 7 A). We could only detect minimal change of the function from linear regression between the dilution assay of the synthetic RNA and the clinical samples (Fig. 7 A, B, C). The slope of the regression function is mainly determined by the chosen number of cycles for the endpoint determination. Compared with the Drosten protocol the CDC panel showed a saturation effect at a grey level of 150 (Fig. 7 D, E).

**Figure 7:**
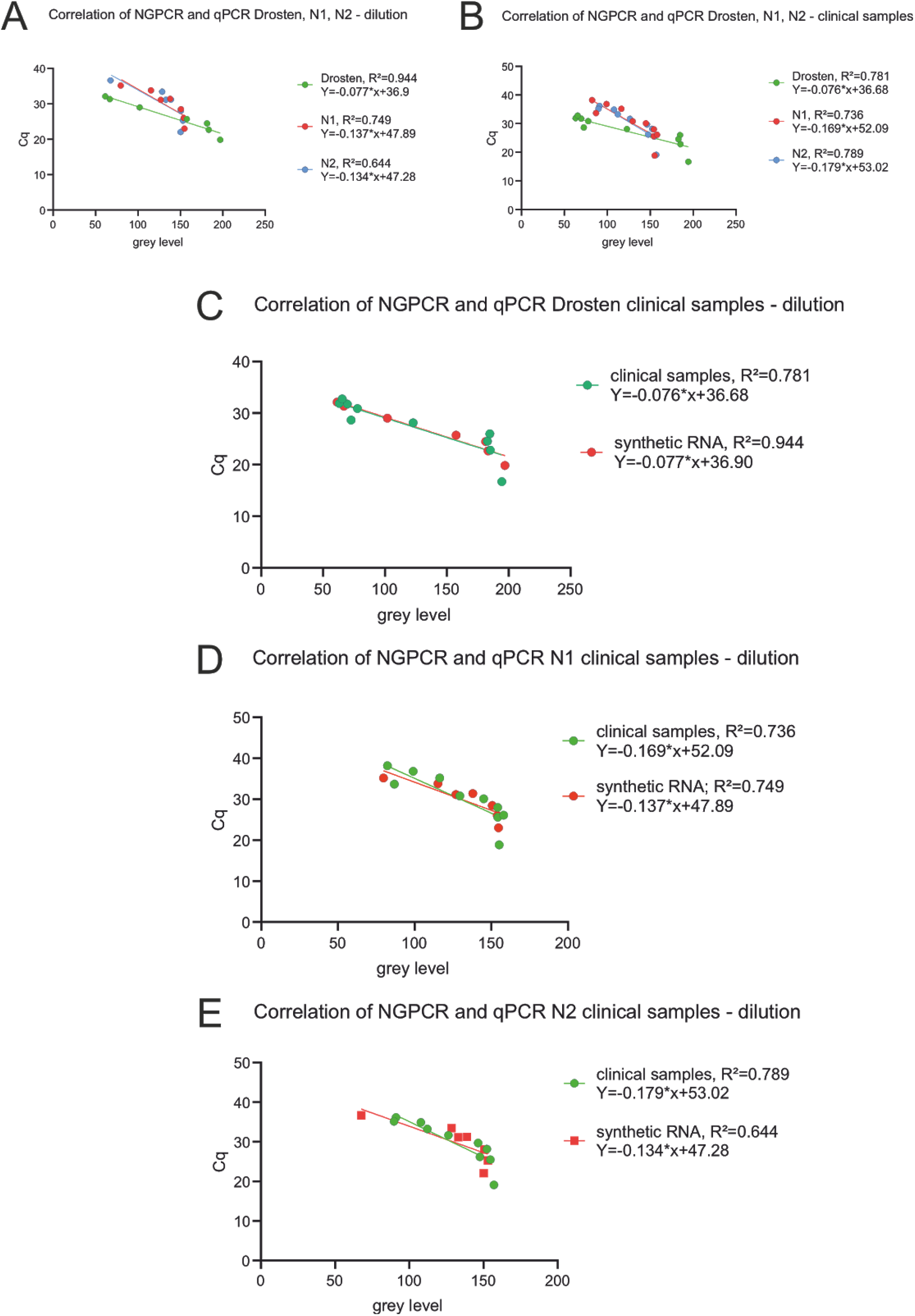
Correlation of Ct values from RT-qPCR and the endpoint fluorescence measurement after NEXTGENPCR: Correlation of NEXTGENPCR and RT-qPCR measurements of synthetic RNA samples with the Drosten-panel (green) as well as with the CDC N1-(red) and N2-protocols (blue) **(A)**. Correlation of NEXTGENPCR and RT-qPCR measurements of clinical RNA samples with the Drosten-panel (green) as well as with the CDC N1-(red) and N2-protocols (blue) **(B)**. Correlation of NEXTGENPCR and RT-qPCR measurements of synthetic RNA (green) and clinical RNA samples (red) with the Drosten-panel **(C)**. Correlation of NEXTGENPCR and RT-qPCR measurements of synthetic RNA (green) and clinical RNA samples (red) with the CDC N1-protocol **(D)**. Correlation of NEXTGENPCR and RT-qPCR measurements of synthetic RNA (green) and clinical RNA samples (red) with the CDC N2-protocol **(E)**.

For the Drosten panel endpoint assay, a trend of saturation can be observed for fluorescence values above 185 (Fig 7 C). This either results from optical limitations of the scanner or from chemical limitations as a component of the chemical reaction might be spent. We hypothesized, that for the CDC protocol saturation effects might have biochemical reasons as it appears earlier than in the Drosten protocol.

To determine whether the saturation effect in the Drosten panel results from optical limitations, a dilution experiment was measured at different scanner exposure time settings. As the spread of the data is maximal between 2x and 3x exposure time and decreases in higher exposure time settings, the observed saturation effect is of optical nature. Of note, different exposure time settings are needed to reach an optimal distinguishability of data points on the lower or higher ends of the spectrum.

As mentioned above, the adaptation of other RT-qPCR based protocols should work in a similar fashion, with the possibility of further cost reduction. At the time of publication, the three other SARS-CoV-2 assays shown in Table 5 have also been optimized on the NEXTGENPCR (GDV, PVH personal communication). Table 5 shows five different RT-qPCR based SARS-CoV-2 test panels with the associated costs. While reagent costs vary between 1,59 and 1,93 € per reaction, the main difference is the performed number of reactions in the protocols. However, the less genes are included in the panel, the higher the possibility of false results.

**Table 5:** Calculated costs for different RT-qPCR based tests. Calculated with the use of VWR nuclease free water and estimated reagent volumes if exact measures weren’t avaiable.

| Creator | Tested region | Total cost per reaction [€] | Total cost per complete panel [€] |
| --- | --- | --- | --- |
| **CDC** | N1 | 1,68 | 5,05 |
|  | N2 | 1,68 |  |
|  | RP | 1,68 |  |
| **Drosten** | E-gene | 1,93 | 5,15 |
|  | RdRP conformation | 1,63 |  |
|  | RdRP discrimination | 1,59 |  |
| **China CDC** | ORF1AB | 1,61 | 3,22 |
|  | N gene | 1,61 |  |
| **Pasteur** | RdRP gene 1 | 1,61 | 5,23 |
|  | RdRP gene 2 | 1,69 |  |
|  | E gene | 1,93 |  |
| **Peckova** | 5-UTR | 1,69 | 1,69 |

## Limitations

The sensitivity was considered using RNA or plasmid standards and eluates of clinical samples. Another important aspect is the preparation of PCR templates (e.g. isolation of nucleic acids), which is not considered in this study. The most obvious limitation of the endpoint PCR is a need for an additional analysis step using the fluorescence scanner. We noted before that the sensitivity of the Drosten panel in the NEXTGENPCR is reduced in low copy number clinical samples (Fig. 4). As shown in Figure 8, the scanner has some optical limitations. Adjusting the scanning settings for low fluorescence and high fluorescence wells separately might optimize sensitivity, while also allowing good correlation with corresponding Cq values. Future effort could be concentrated on the development of better fluorescence scanners and or the development of a real time extension for NGPCR. In its current version NEXTGENPCR is fast and easy to use, and hands on time is reduced when integrated into a robotic workflow.

**Figure 8:**
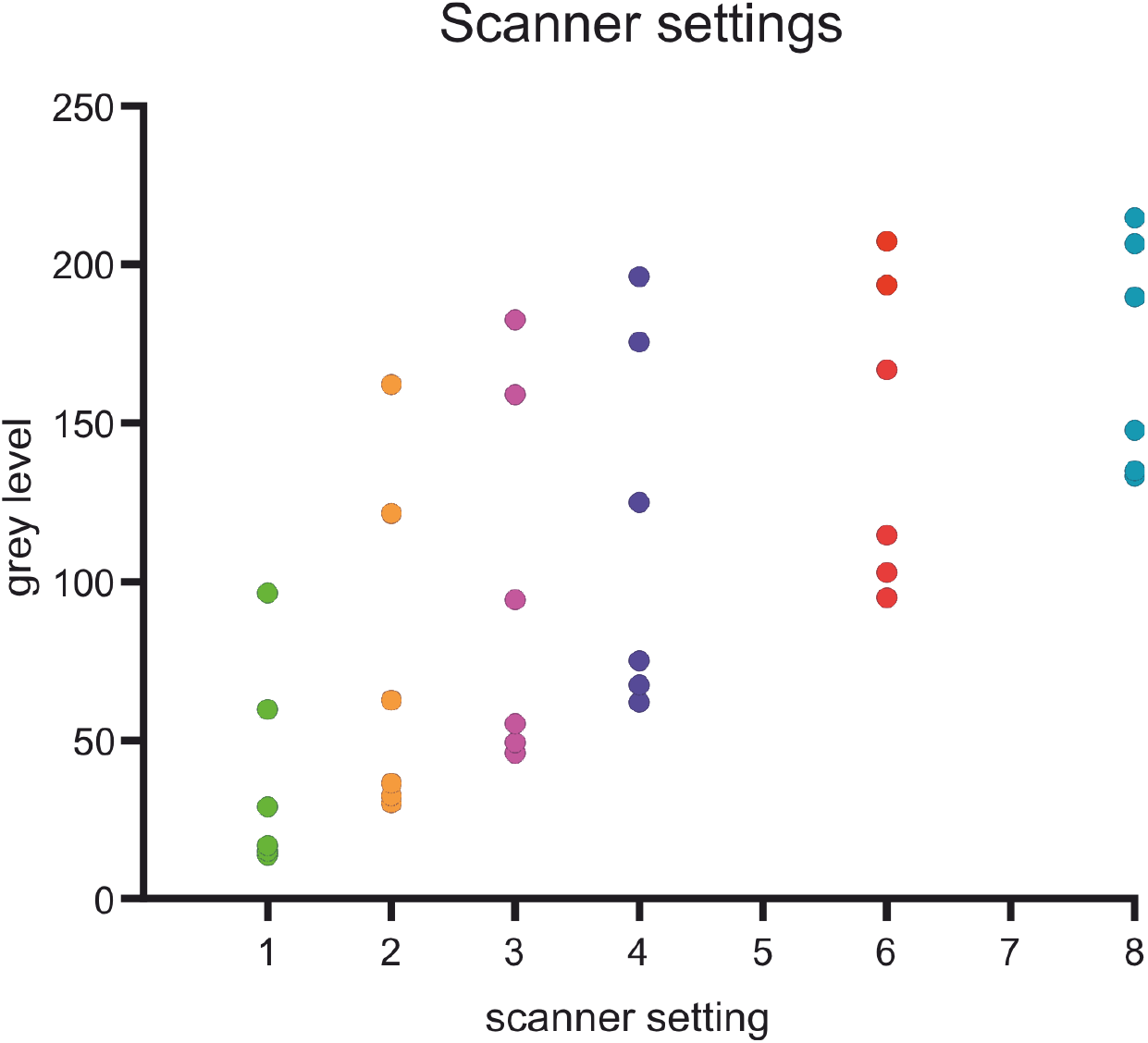
Optical limitations of the scanning system: Measurements of fluorescence signals after NEXTGENPCR of synthetic RNA samples with scanner settings of 1x, 2x, 3x, 4x, 6x and 8x amplification.

## Conclusions

Although usually qPCR is used to score and quantify the amount of virus in a sample, it is not clear that the amount of sample is related to the severity of the disease. Rather certain genetic properties could be linked to the severity of the disease. Furthermore, RT-qPCR platforms have higher costs and rely on highly skilled personnel. Therefore, it is only logical to reduce the complexity of the method of detection and rely on endpoint PCR. Using a NEXTGENPCR machine and RNA extraction rather than isolation brings a complete test from sample to result in less than 30 minutes into view. The method described here would facilitate both population screening or point-of-care testing.

## Data Availability

All data generated or analyzed during this study are included in this article or can be requested by the author for correspondence: Christian Kaltschmidt

## ACKNOWLEDGEMENTS

We acknowledge funding of the University of Bielefeld and a supply of reagents and consumables from Molecular Biology Systems B.V., Goes, The Netherlands.

